# External Validation of an Automated Segmentation Tool for Abdominal Fat Tissue using DIXON MRI: Data from the CutDM trial

**DOI:** 10.1101/2025.05.14.25327583

**Authors:** Frederik Hvid Linden, Mikael Boesen, Erik Høegh-schmidt, Janus Uhd Nybing, Rasmus Bastkjær Mainz Hansen, Luise Helene Persson Kopp, Mads Norvin Thomsen, Mathias Willadsen Brejnebøl

**Affiliations:** Department of Radiology, Bispebjerg and Frederiksberg Hospital, Copenhagen, Denmark; Department of Endocrinology, Copenhagen University Hospital Bispebjerg, Copenhagen, Denmark

## Abstract

1.

**Background and rationale:** Segmentation and quantifying the volume of visceral adipose tissue (VAT) and subcutaneous adipose tissue (SAT) is an emerging field that has shown to be highly relevant for studying the risk of various metabolic diseases and monitoring treatment effects over time. The reference method for segmenting VAT and SAT involves manual segmentation in a prespecified volume, such as delimited by the cephalic part of L3 to the caudal part of L5 of the lumbar spine using the fat image produced by the DIXON technique. However, scalability and more widespread use is challenged by the time-consuming nature and potential inter- and intrareader variability of the manual approach.

Lately increasing number of segmentation tools have been made available under open source such as the FatSegNet. For such tools, only the internal diagnostic test accuracy is reported. Furthermore, segmentation tools rarely have a proper localizer which is required for an end-to-end workflow.

**Objectives:** We will prepend the existing FatSegNet model with a L3 to L5 delimiter model to establish a pipeline for fully automated analysis of an axial Dixon stack. The aim of this study will be to test the diagnostic accuracy this full VAT-SAT quantification approach.

**Methods:** Two experienced readers will manually segment the VAT and the SAT for each slice of the abdominal volume defined by the axial slice through the cephalic part of the L3 vertebra to caudal part of the L5 vertebra in 32 participants (16 male and 16 female) with BMI >25 who underwent 6-point mDIXON QUANT (Philips Healthcare) at baseline and follow up.

**Population:** The sample for this study inherits the characteristics of the CUTDm trial with the eligibility criteria:

- Diabetes mellitus type 2
- BMI > 25
- Men or postmenopausal women aged 18-75 years. Menopause will be defined as >12 months without menses
- HbA1c 48-75 mmol/mol (6.5%-9.0%)
- Treated with or without Metformin, DPP-4 inhibitors, SGLT-2 inhibitors and/or GLP-1 receptor agonists (GLP-1RA)
- Non-smokers or having quitted smoking >1 year before inclusion in the study
- Acceptance of regulation of antidiabetic, antihypertensive, and lipid-lowering medications by the Cut-DM endocrinologists only

**Index test:** The segmented volumes for VAT and SAT outputted by the AI tool.

**Reference test:** The scan-level mean segmented volumes of the two independent, reference readers.

*Further statistical details:* 

*Sample size:* Not applicable as this is a secondary analysis.

*Framework:* 

*Confidence intervals and P values:* All 95% confidence intervals.

*Multiplicity:* No explicit multiplicity correction will be performed.

*Statistical software:* R version 4.2.2 (or newer).

## 2 ELABORATIONS ON OUTCOMES AND DATA

### Data management

The volumes segmented by the AI tool will be compared to the average segmented volumes of two independent experienced readers.

#### SAT

Segmentation of the subcutaneous abdominal fat. Cm3.

#### VAT

Segmentation of the visceral abdominal fat. Cm3.

#### Change in SAT & VAT

Changes in abdominal fat volumes of participants from baseline to one year follow up.

### Data validation

All variables used in the analyses, including the derived variables, will be checked for missing values, outliers, and inconsistencies.

#### Data template

Based on this SAP, the statistical analyst will develop a tailored data template illustrating the data structure required for the statistical analyses.

## 3 OUTLINE

The analysis success rate of the AI tool (defined as the number of patient scans that the AI tool successfully analysed, unrelated to the actual segmented volume), and the agreement between the independent segmentations of the reference readers will only be reported in the manuscript body.

The anticipated (predefined) outline of the manuscript is illustrated below.

### Further statistical information related to Table 1

Data will be presented as means with standard deviations (SD) when normally distributed or as medians with interquartile range in case of skewed data. Dichotomous and categorical data will be presented as absolute counts and proportions.

**Table 1.** Patient Characteristics.

| Characteristic | Total<br>(n = x) |
| --- | --- |
| Female |  |
| Male |  |
| Age (y) |  |
| Age range (y) |  |
| BMI(kg/m<sup>2</sup>) |  |
| SAT(cm<sup>3</sup> SD) |  |
| VAT(cm<sup>3</sup>) |  |

**Table 2.**
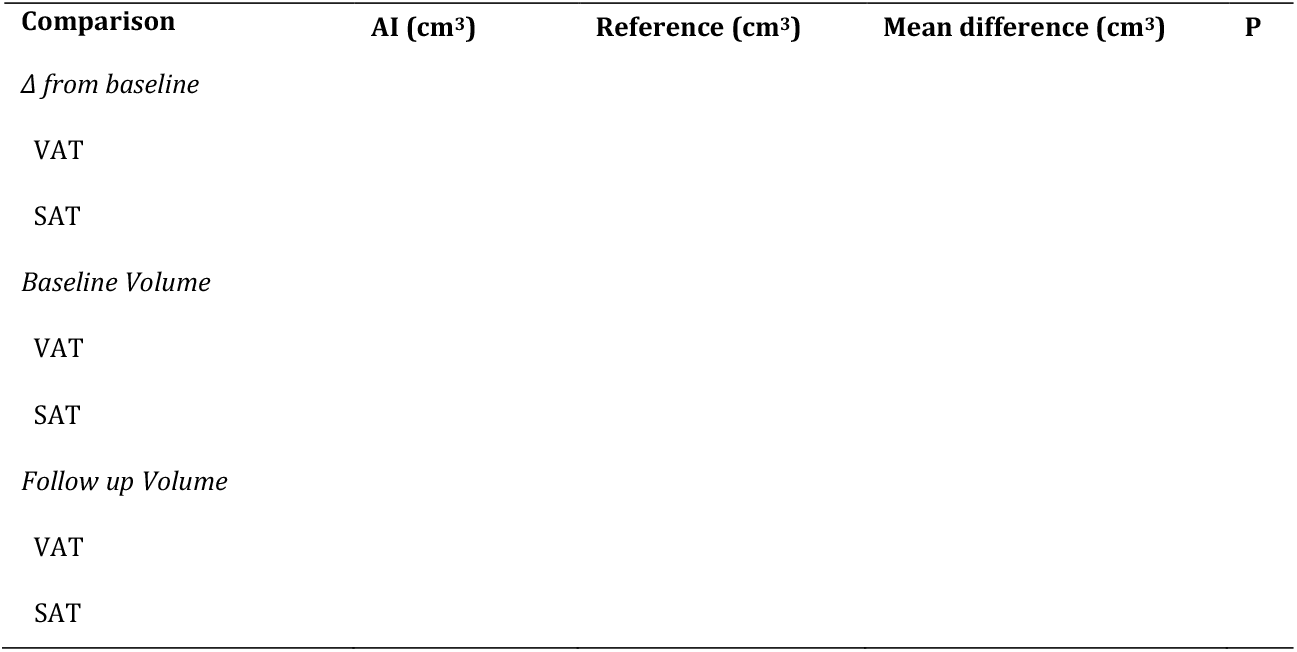
Diagnostic test accuracy of the AI tool. compared to the mean segmented volume of the reference readers. The P-value column signifies the P-value of the two-tailed student’s *t*-test.

### Hypothetical box and whisker plot for the AI tool and for the readers

After consulting the literature and international experts in the field, it has not been possible to find a specific threshold for the increase in VAT and SAT in relation to the risk of developing diabetes and other metabolic diseases. In this study, equivalence of the AI tool is established if the bounds of the 95% confidence interval for the AI tools segmented volume for the change from baseline to follow-up is within 10% of the reference. (1) (2) (3).

### Further statistical information related to Figure 2

The **box and whisker plot** visually represents the diagnostic test accuracy of the AI tool. The **red line** denotes the **median differences**, the **yellow box** represents the **standard deviation**, and the whiskers extend to 1.5 times the interquartile range (IQR). This mockup plot demonstrates the various ways the AI tool ends up performing.

**Figure 1.**
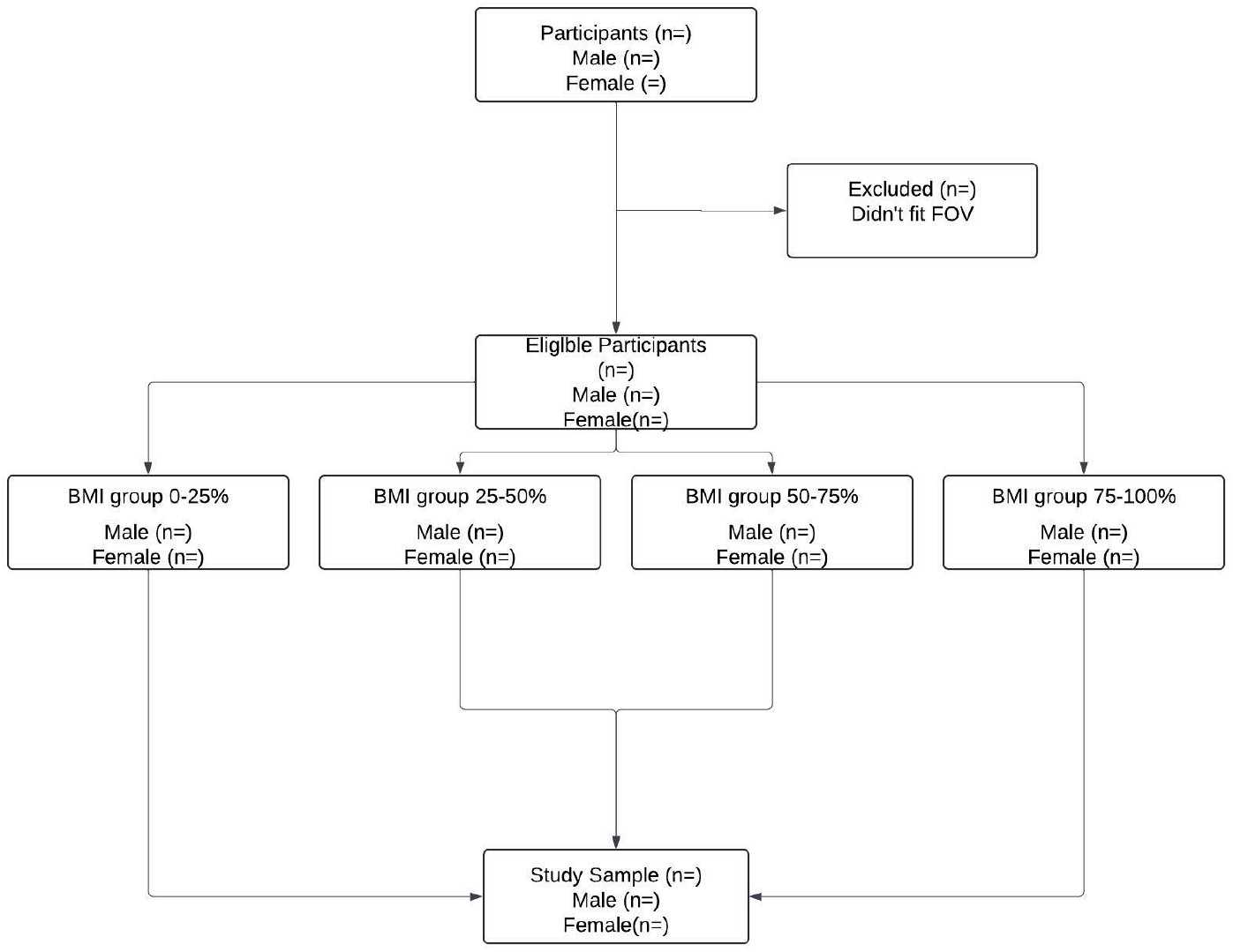
Flow diagram. Anticipated plot design, illustrating potential reasons for exclusion:

**Figure 2.**
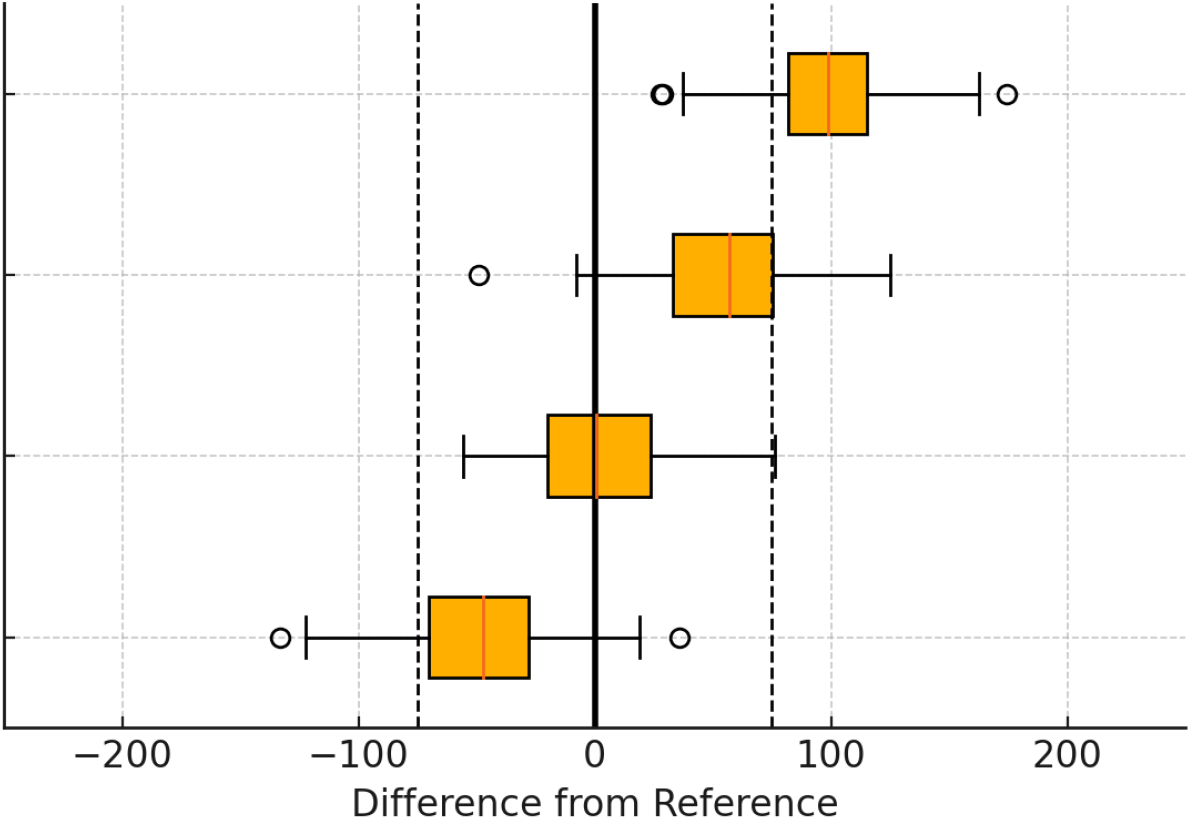
Difference between the AI tool and the reference

*Hypothetical Bland-Altman plot of the independently segmented volumes of the reference readers*

### Further statistical information related to Figure 3

Standard 95% limits of agreement will be used for the Bland-Altman plot.

**Figure 3.**
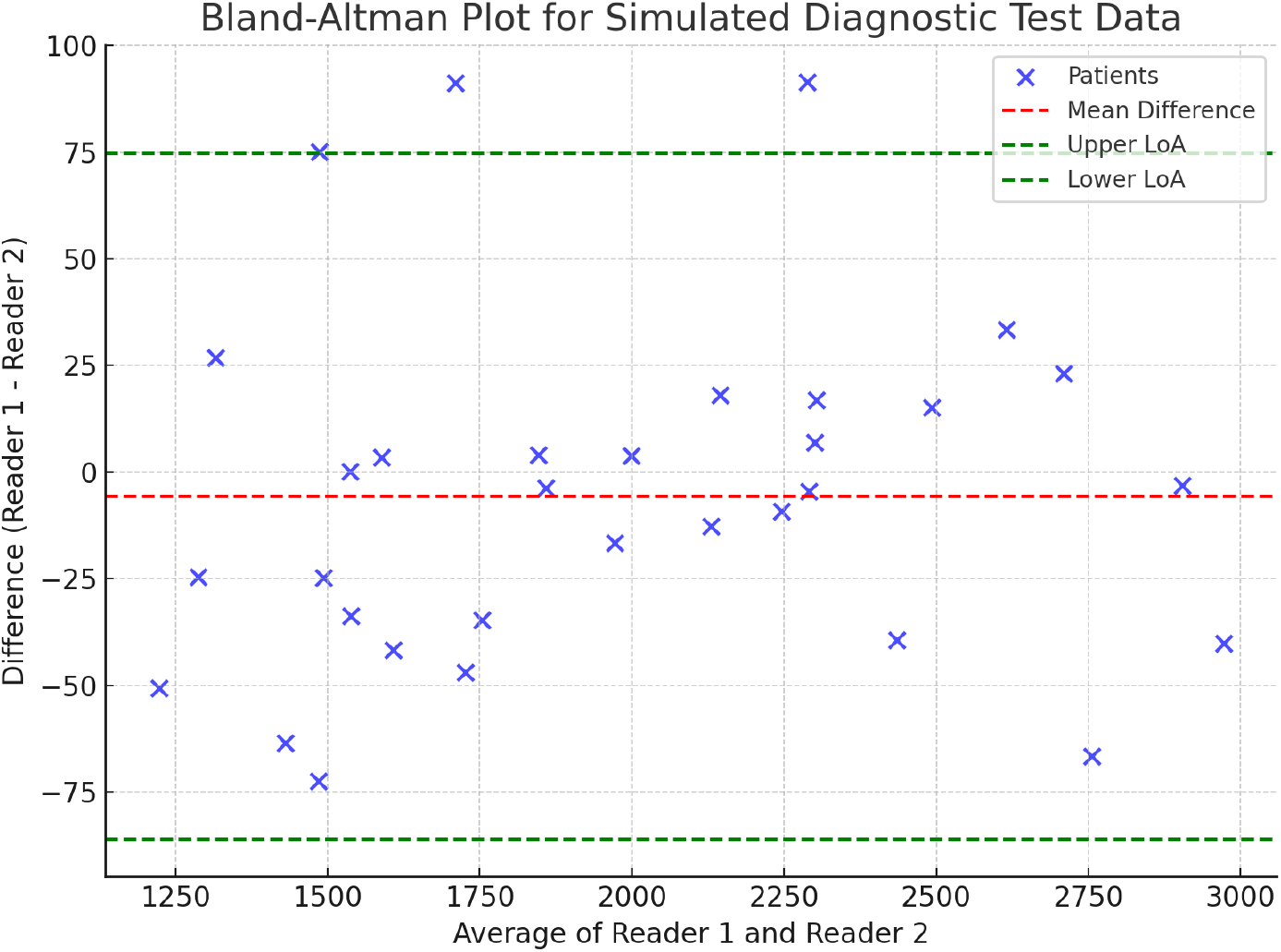
Bland-Altman plot of the reference readers (Mock up)

Same as the primary analysis (Table 3) but divided in different BMI levels. Values will be reported as the AUC (95% confidence interval). Readers will refer to the two independent, experienced reference readers.

**Table 3.**
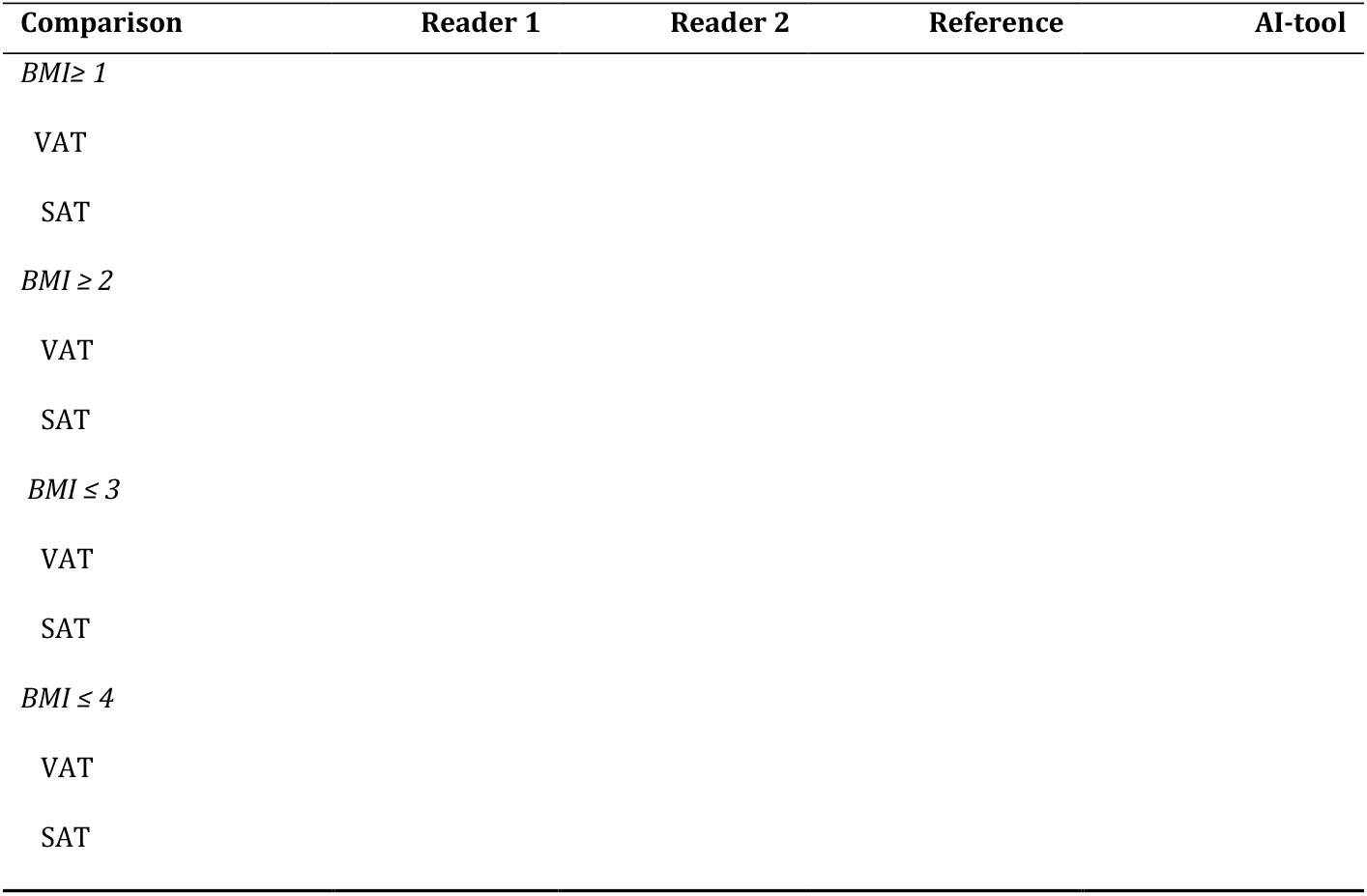
Diagnostic test accuracy of the AI tool for SAT and VAT at different BMI quartiles.

## Data Availability

All data produced in the present study are available upon reasonable request to the authors

## SUPPLEMENTARY MATERIAL

The anticipated (predefined) supplementary material of the manuscript is illustrated below.

### Supplementary file 1. Protocol[1]

### Supplementary file 2. This SAP

**Supplementary Table 1.**
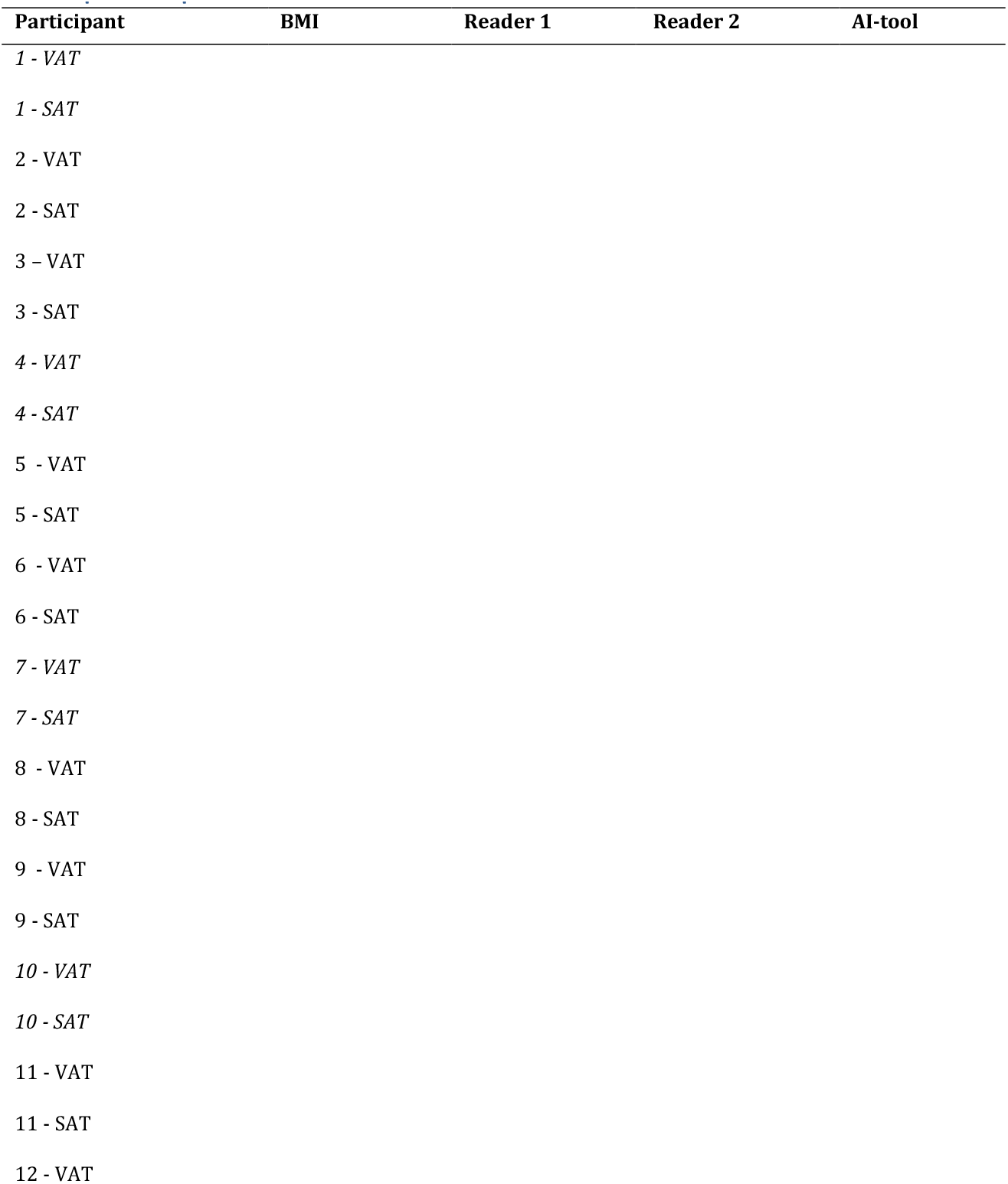
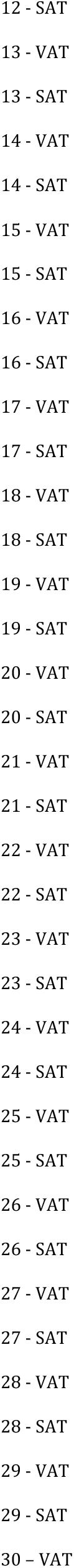
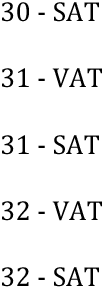
Raw segmented values for VAT and SAT for each participant

## 5. SAP REPORTING GUIDELINE

This SAP has been reported according to the items recommended in STARD guidelines by Cohen et al.[7] Explanation and elaboration of the items are available in the appendix paper.[8]

The guideline checklist and motivation is reproduced below.

| Section & Topic | No | Item | Reported on page # |
| --- | --- | --- | --- |
| TITLE OR ABSTRACT |  |  |  |
|  | 1 | Identification as a study of diagnostic accuracy using at least one measure of accuracy<br>(such as sensitivity, specificity, predictive values, or AUC) |  |
| ABSTRACT |  |  |  |
|  | 2 | Structured summary of study design, methods, results, and conclusions<br>(for specific guidance, see STARD for Abstracts) |  |
| INTRODUCTION |  |  |  |
|  | 3 | Scientific and clinical background, including the intended use and clinical role of the index test |  |
|  | 4 | Study objectives and hypotheses |  |
| **METHODS** |  |  |  |
| *Study design* | 5 | Whether data collection was planned before the index test and reference standard were performed (prospective study) or after (retrospective study) |  |
| *Participants* | 6 | Eligibility criteria |  |
|  | 7 | On what basis potentially eligible participants were identified (such as symptoms, results from previous tests, inclusion in registry) |  |
|  | 8 | Where and when potentially eligible participants were identified (setting, location and dates) |  |
|  | 9 | Whether participants formed a consecutive, random or convenience series |  |
| *Test methods* | 10a | Index test, in sufficient detail to allow replication |  |
|  | 10b | Reference standard, in sufficient detail to allow replication |  |
|  | 11 | Rationale for choosing the reference standard (if alternatives exist) |  |
|  | 12a | Definition of and rationale for test positivity cut-offs or result categories of the index test, distinguishing pre-specified from exploratory |  |
|  | 12b | Definition of and rationale for test positivity cut-offs or result categories of the reference standard, distinguishing pre-specified from exploratory |  |
|  | 13a | Whether clinical information and reference standard results were available to the performers/readers of the index test |  |
|  | 13b | Whether clinical information and index test results were available to the assessors of the reference standard |  |
| *Analysis* | 14 | Methods for estimating or comparing measures of diagnostic accuracy |  |
|  | 15 | How indeterminate index test or reference standard results were handled |  |
|  | 16 | How missing data on the index test and reference standard were handled |  |
|  | 17 | Any analyses of variability in diagnostic accuracy, distinguishing pre-specified from exploratory |  |
|  | 18 | Intended sample size and how it was determined |  |
| **RESULTS** |  |  |  |
| *Participants* | 19 | Flow of participants, using a diagram |  |
|  | 20 | Baseline demographic and clinical characteristics of participants |  |
|  | 21a | Distribution of severity of disease in those with the target condition |  |
|  | 21b | Distribution of alternative diagnoses in those without the target condition |  |
|  | 22 | Time interval and any clinical interventions between index test and reference standard |  |
| *Test results* | 23 | Cross tabulation of the index test results (or their distribution) by the results of the reference standard |  |
|  | 24 | Estimates of diagnostic accuracy and their precision (such as 95% confidence intervals) |  |
|  | 25 | Any adverse events from performing the index test or the reference standard |  |
| **DISCUSSION** |  |  |  |
|  | 26 | Study limitations, including sources of potential bias, statistical uncertainty, and generalisability |  |
|  | 27 | Implications for practice, including the intended use and clinical role of the index test |  |
| **OTHER INFORMATION** |  |  |  |
|  | 28 | Registration number and name of registry |  |
|  | 29 | Where the full study protocol can be accessed |  |
|  | 30 | Sources of funding and other support; role of funders |  |

## STARD 2015

### AIM

STARD stands for “Standards for Reporting Diagnostic accuracy studies”. This list of items was developed to contribute to the completeness and transparency of reporting of diagnostic accuracy studies. Authors can use the list to write informative study reports. Editors and peer-reviewers can use it to evaluate whether the information has been included in manuscripts submitted for publication.

### Explanation

A **diagnostic accuracy study** evaluates the ability of one or more medical tests to correctly classify study participants as having a **target condition**. This can be a disease, a disease stage, response or benefit from therapy, or an event or condition in the future. A medical test can be an imaging procedure, a laboratory test, elements from history and physical examination, a combination of these, or any other method for collecting information about the current health status of a patient.

The test whose accuracy is evaluated is called **index test**. A study can evaluate the accuracy of one or more index tests. Evaluating the ability of a medical test to correctly classify patients is typically done by comparing the distribution of the index test results with those of the **reference standard**. The reference standard is the best available method for establishing the presence or absence of the target condition. An accuracy study can rely on one or more reference standards.

If test results are categorized as either positive or negative, the cross tabulation of the index test results against those of the reference standard can be used to estimate the **sensitivity** of the index test (the proportion of participants *with* the target condition who have a positive index test), and its **specificity** (the proportion *without* the target condition who have a negative index test). From this cross tabulation (sometimes referred to as the contingency or “2×2” table), several other accuracy statistics can be estimated, such as the positive and negative **predictive values** of the test.

Confidence intervals around estimates of accuracy can then be calculated to quantify the statistical **precision** of the measurements.

If the index test results can take more than two values, categorization of test results as positive or negative requires a **test positivity cut-off**. When multiple such cut-offs can be defined, authors can report a receiver operating characteristic (ROC) curve which graphically represents the combination of sensitivity and specificity for each possible test positivity cut-off. The **area under the ROC curve** informs in a single numerical value about the overall diagnostic accuracy of the index test.

The **intended use** of a medical test can be diagnosis, screening, staging, monitoring, surveillance, prediction or prognosis. The **clinical role** of a test explains its position relative to existing tests in the clinical pathway. A replacement test, for example, replaces an existing test. A triage test is used before an existing test; an add-on test is used after an existing test.

Besides diagnostic accuracy, several other outcomes and statistics may be relevant in the evaluation of medical tests. Medical tests can also be used to classify patients for purposes other than diagnosis, such as staging or prognosis. The STARD list was not explicitly developed for these other outcomes, statistics, and study types, although most STARD items would still apply.

### DEVELOPMENT

This STARD list was released in 2015. The 30 items were identified by an international expert group of methodologists, researchers, and editors. The guiding principle in the development of STARD was to select items that, when reported, would help readers to judge the potential for bias in the study, to appraise the applicability of the study findings and the validity of conclusions and recommendations. The list represents an update of the first version, which was published in 2003.

More information can be found on http://www.equator-network.org/reporting-guidelines/stard.

